# Cardiovascular Risk After Cancer in Patients with Ischemic Heart or Peripheral Artery Disease

**DOI:** 10.64898/2025.12.04.25341674

**Authors:** Torben Bjerregaard Larsen, Ricco Noel Hansen Flyckt, Margrethe Bang Henriksen, Claus Lohman Brasen, Torben Frøstrup Hansen, Flemming Skjøth

**Author notes:** **Address for correspondence:** Professor Torben Bjerregaard Larsen, Beriderbakken 4, DK-7100 Vejle, Denmark. **Tweet:** “In >91,000 Danish patients with IHD/PAD who develop cancer, 1 year mortality is 33% and CV events are frequent. Early integrated cardio oncology care and aggressive risk factor control may improve outcomes. #cardiooncology #CVD #cancer”.

## Abstract

**Objective:** To quantify 1-year risks of cardiovascular death, major adverse cardiovascular events (MACE), and venous thromboembolism (VTE) after cancer diagnosis in patients with pre-existing ischemic heart disease (IHD) or peripheral artery disease (PAD), and to identify clinical predictors of early vascular events in this high-risk population.

**Approach and Results:** Using nationwide Danish registries, we identified 91,253 patients with IHD and/or PAD who developed a first primary cancer between 2002 and 2022. Competing-risk methods were used to estimate 1-year cumulative incidences of cardiovascular outcomes, and multivariable Cox models assessed associations with baseline clinical predictors.

One-year cumulative incidences were 2.4 percent for cardiovascular death, 2.8 percent for MACE, and 1.8 percent for VTE, with substantial variation across cancer types. Neurologic, respiratory, and digestive cancers showed the highest vascular event rates. Prior stroke, myocardial infarction, and higher comorbidity burden were strong predictors of MACE, while previous VTE strongly predicted recurrent thrombosis. Statin therapy was associated with lower cardiovascular mortality (HR 0.66) and fewer MACE events (HR 0.84).

**Conclusion:** Cancer diagnosis in patients with established atherosclerotic disease is followed by a substantial burden of early cardiovascular and thrombotic events. Distinct risk profiles across malignancy types and the protective association of statins highlight the relevance of vascular biology in shaping early outcomes and support continued use of guideline-directed preventive therapies during cancer care.

## Introduction

Ischemic heart disease (IHD) and peripheral artery disease (PAD) are serious manifestations of systemic atherosclerosis and are associated with high rates of morbidity and mortality.^1^ Many patients with IHD or PAD require intensive cardiovascular care, including anticoagulation, revascularization procedures, and commonly present with additional comorbidities such as diabetes, heart failure, and atrial fibrillation. Cancer is also prevalent in this population, driven by shared risk factors including older age, smoking, and chronic inflammation.^2^ Patients with IHD or PAD represent an advanced vascular phenotype characterized by endothelial dysfunction, chronic inflammation, and heightened thrombotic potential. These biological features may be amplified during cancer development through systemic inflammatory activation, coagulation pathway stimulation, and cancer-associated metabolic stress.^2–4^ Consequently, the vascular system is directly challenged when cancer arises in patients with existing atherosclerotic disease, but the magnitude and timing of early cardiovascular complications in this phenotype remain poorly quantified in population-based settings.

Although cancer and cardiovascular disease are typically managed along separate clinical pathways, their interaction is increasingly recognized. Cancer therapies can accelerate cardiovascular injury, while underlying vascular disease may limit treatment options or increase the risk of toxicity. Patients with IHD or PAD who develop cancer represent a particularly vulnerable subgroup, often excluded from clinical trials and underrepresented in current cardio-oncology guidelines.^5^

Accurate cardiovascular risk stratification is critical in oncology to guide surveillance and inform treatment decisions. Previous studies have shown that prediction models based on routinely collected registry data may be feasible and clinically informative. Applying such models in cancer populations with pre-existing atherosclerotic disease could help personalize and improve cardio-oncological care.^6,7^

Understanding vascular risk in this population is essential not only for clinical management but also for clarifying how cancer-related biological stressors interact with pre-existing atherosclerotic disease and thrombotic pathways. In this study, we use nationwide registry data to analyze a large population of Danish patients with IHD and/or PAD who subsequently developed cancer. We describe rates of all-cause mortality and major adverse cardiovascular events (MACE) and evaluate clinical predictors with the goal of assessing the potential for registry-based risk modeling in this complex patient population.

## Methods

### Data Sources

This study used data from four nationwide Danish health registries, all linked at the individual level through a unique personal identification number assigned to every Danish resident at birth or upon immigration.^8^ These registries offer comprehensive longitudinal data on demographics, comorbidities, healthcare utilization, and mortality.

The Danish National Patient Register, established in 1977, includes diagnostic codes and records of medical procedures from all somatic hospital admissions.^8^ We classified diagnoses using the International Classification of Diseases, 10th revision (ICD-10), which has been implemented in Denmark since 1997. The Danish National Prescription Registry contains data on all redeemed prescriptions since 1994 and was used to identify pharmacologic exposures.^9^ We categorized pharmacologic treatments according to the Anatomical Therapeutic Chemical (ATC) Classification System.

Demographic data—including date of birth, sex, migration status, and vital status—were obtained from the Danish Civil Registration System.^8^ Information on cause-specific mortality was retrieved from the Danish National Cause of Death Register, which was available since 2002.^10^

Access to the data used in this study was granted through the Open Patient data Explorative Network (OPEN), Odense University Hospital, Region of Southern Denmark, and Statistics Denmark. The study was approved by the Danish Data Protection Agency via institutional registration with the Region of Southern Denmark (reference 23-50300). Access to Danish health registry data requires affiliation with, or a formal collaboration involving, a Danish research institution as well as prior authorization from the relevant data custodians. All data are stored and analyzed within secure research environments administered by Statistics Denmark and cannot be transferred, downloaded, or exported for external use.

### Study Population and Outcomes

Cardiovascular outcomes were selected to reflect biologically linked processes involving atherosclerotic plaque instability (MACE), thrombotic events (VTE), and vascular mortality pathways.

We identified all patients with a hospital discharge diagnosis of ischemic heart disease (IHD; ICD-10 codes I20–I25) or peripheral artery disease (PAD; ICD-10 codes I70–I74, I77), excluding subcodes representing other peripheral vascular conditions (I708A, I713, I714, I730, I731, I738, I739B). Patients were included if they subsequently received a diagnosis of cancer (ICD-10 codes C00–C97) between January 1, 2002, and June 30, 2022.

Patients with ICD-10 code C44 (“Other and unspecified malignant neoplasms of skin”) were excluded. This code captures nonmelanoma skin cancers, including basal cell carcinoma and squamous cell carcinoma, but excludes melanoma, which is classified under code C43.

The date of cancer diagnosis was defined as the index date for follow-up.

To ensure data quality and identify incident cases, patients with inconsistent vital status information or less than 1 year of residence in Denmark prior to cancer diagnosis were excluded. Individuals with a history of cancer preceding the diagnosis of IHD or PAD were also excluded, as were those diagnosed with unspecified or metastatic cancers (ICD-10 codes C76–C80).To account for variation in cancer prognosis, malignancies were categorized by anatomic site: digestive tract (C00–C26); respiratory organs, including intrathoracic structures (ICD-10 codes C30–C39); bone, skin, and connective tissue (C40–C49); glandular and reproductive organs (C50–C63, C73–C75); uro-renal system (C64–C68); neurologic system (C69–C72); and hematologic malignancies (C81–C99). These groupings reflect common anatomic divisions and broadly similar treatment considerations within each category.

Primary outcomes were MACE, VTE and cardiovascular death. Cardiovascular death was identified using the Danish Cause of Death Register. MACE was defined as ischemic stroke, systemic embolism, myocardial infarction, or hemorrhagic stroke. Only primary discharge diagnoses from acute hospital encounters were included to avoid misclassifying follow-up visits for prior cardiovascular events as incident events. VTE included pulmonary embolism and deep vein thrombosis. To improve diagnostic specificity, VTE events were included only if the diagnosis was recorded within 10 days of a procedure code indicating imaging or diagnostic evaluation for emboli in the lungs or lower extremities.^11^ Diagnostic codes used to define each outcome are listed in Supplementary Table 1.

Baseline cardiovascular risk factors were assessed using routinely collected administrative data commonly applied in clinical risk prediction models. At the index date, we recorded diagnoses of atrial fibrillation, ischemic stroke, VTE, major bleeding, myocardial infarction, kidney disease, liver disease, heart failure, diabetes mellitus, hypertension, IHD, PAD, rheumatic disease or systemic lupus erythematosus, and thyroid dysfunction (hypo- or hyperthyroidism). Diagnoses of hypertension and diabetes mellitus required evidence of pharmacologic treatment.

Patients were classified as receiving cardiovascular prophylaxis if they filled at least 1 prescription for a statin, antiplatelet agent, or oral anticoagulant in the year before their cancer diagnosis. Overall comorbidity burden was assessed using the Charlson Comorbidity Index, categorized as 0–3, 4, 5, 6, or 7–21 points.^12^ To estimate stroke and cardiovascular risk, we calculated the CHA₂DS₂-VASc score.^13^ The full list of codes used to define risk factors is provided in Supplementary Table 1.

### Statistical Analyses

Time-to-event methods were used to estimate the risk of outcomes and examine associations with cardiovascular risk factors. Time at risk was defined as the period from the cancer diagnosis date (index date) to the earliest of the following: outcome occurrence, death, emigration, 1 year of follow-up, or December 31, 2022.

Crude incidence rates were calculated as the number of events per person-years at risk, reported both overall and stratified by cancer site. To account for competing risks, cumulative incidence functions for MACE and VTE were estimated using the Aalen–Johansen estimator, with all-cause death treated as a competing event. For cardiovascular death, non-cardiovascular deaths were considered competing events. Given the high mortality in this population, we also reported all-cause mortality rates and estimated survival using the Kaplan–Meier method.

Associations between outcomes and clinical predictors were assessed using Cox proportional hazards regression. Models included sex, cancer site, age (modeled as a continuous variable using cubic splines), Charlson Comorbidity Index group, and all cardiovascular risk indicators defined above. Both crude and adjusted hazard ratios (HRs) were reported, with adjustment based on inclusion of all listed predictors in the multivariable model.

All data management and statistical analyses were performed using SAS version 9.4 (SAS Institute Inc), Stata/MP version 18.0 (StataCorp LLC), and R Statistical Software version 4.3.0 (R Foundation for Statistical Computing).

## Results

We initially identified 126,735 patients with IHD and/or PAD who were diagnosed with a first primary cancer between 2002 and 2022. After applying exclusion criteria based on registry data and timing of diagnoses, 91,253 patients were included in the final study population.

The inclusion process is detailed in the study flowchart (Figure 1), which outlines registry definitions, exclusion criteria, and cohort construction.

**Figure 1.**
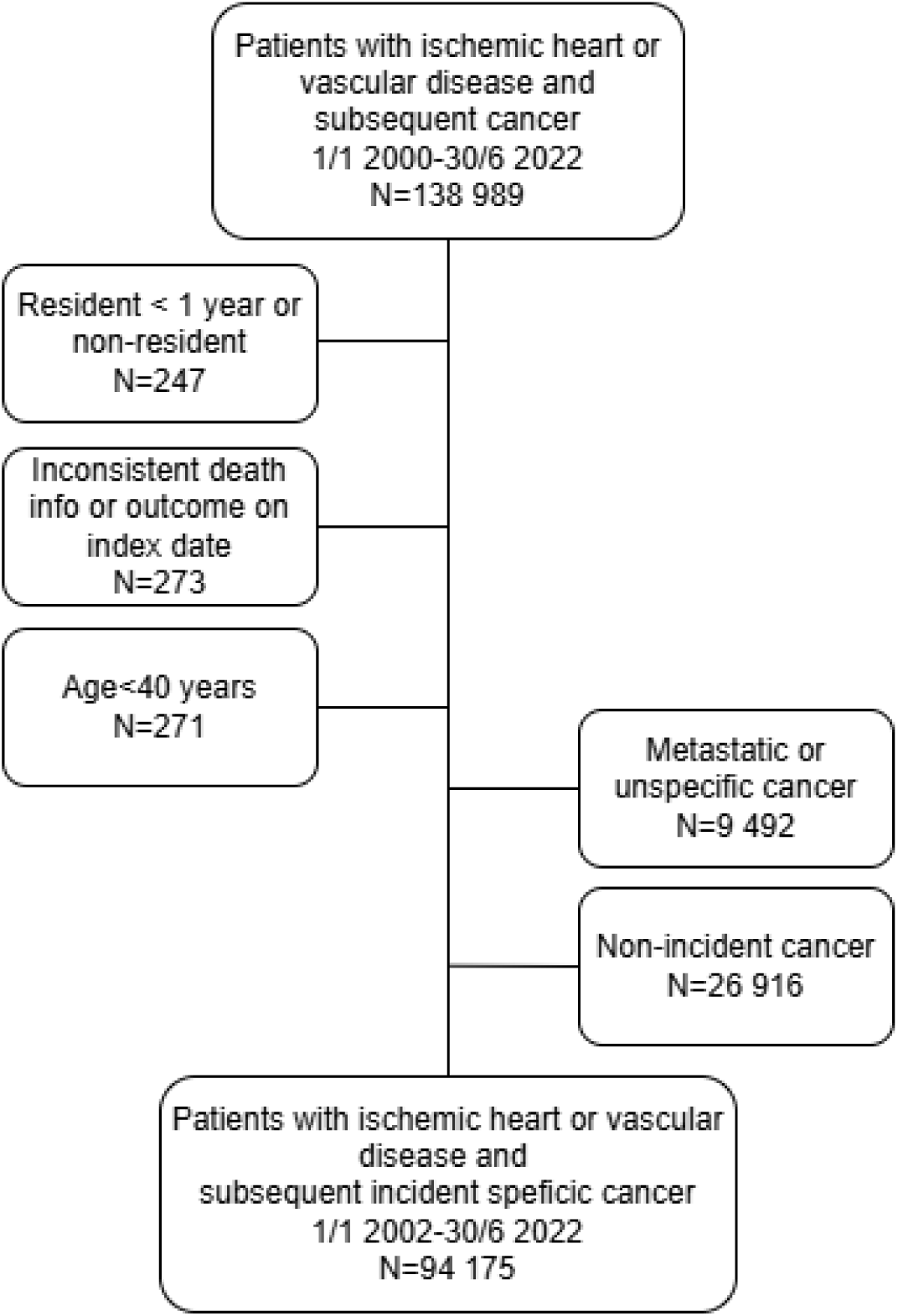
Inclusion of patients. *PAD: Peripheral artery disease; VTE: Venous thromboembolism; CHA₂DS₂-VASc Score: congestive heart failure, hypertension, age ≥75 years (doubled), diabetes, stroke/transient ischemic attack/thromboembolism (doubled), vascular disease, age 65–74 years, female*.

### Baseline Characteristics

Baseline characteristics of the study population are shown in Table 1. The mean age was 73.7 years (SD, 9.6), and 36.7% were female. Ischemic heart disease was the primary inclusion diagnosis in 79.3% of patients, 32.7% had PAD, and 12.0% had both conditions.

**Table 1.**
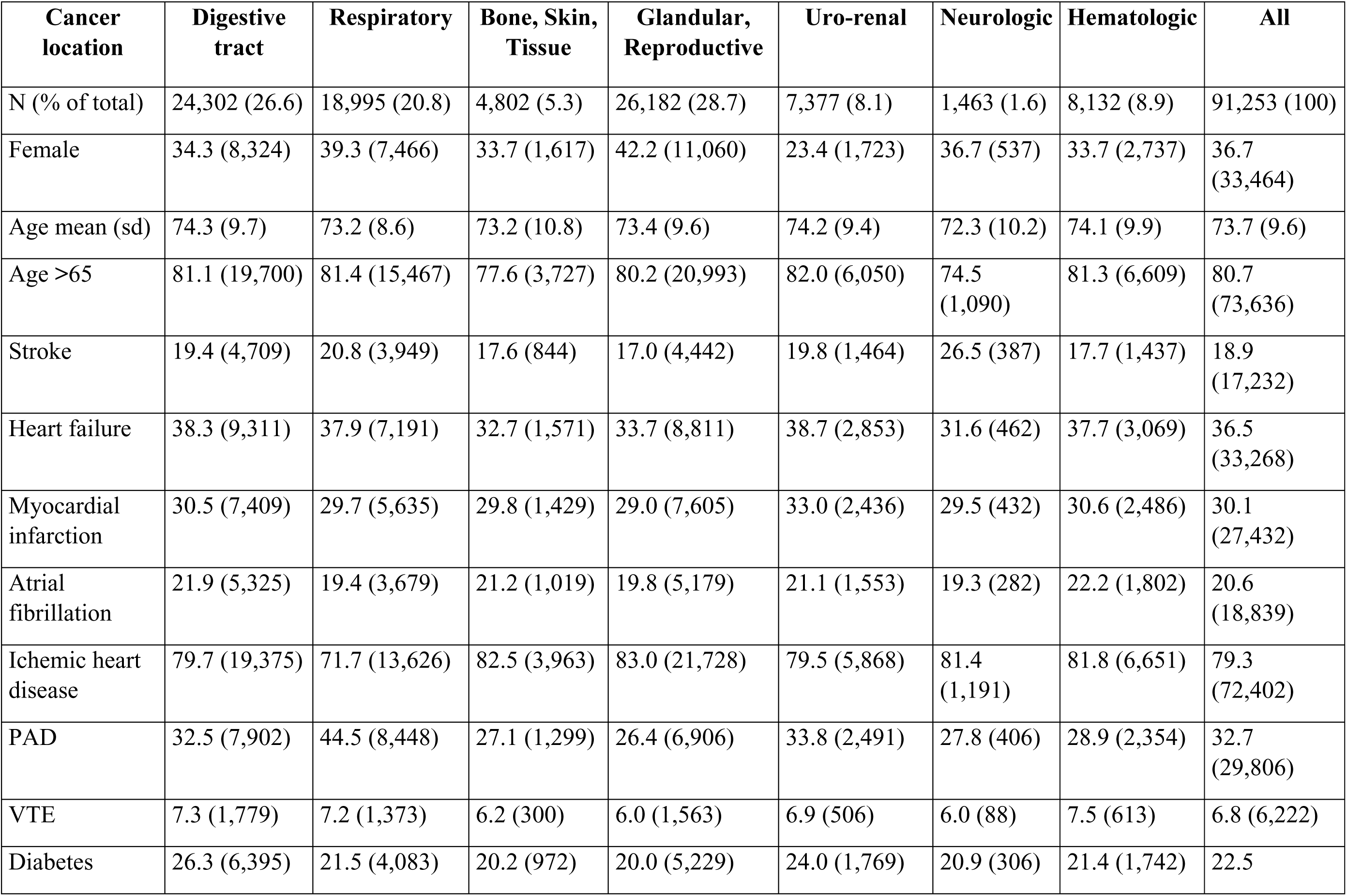
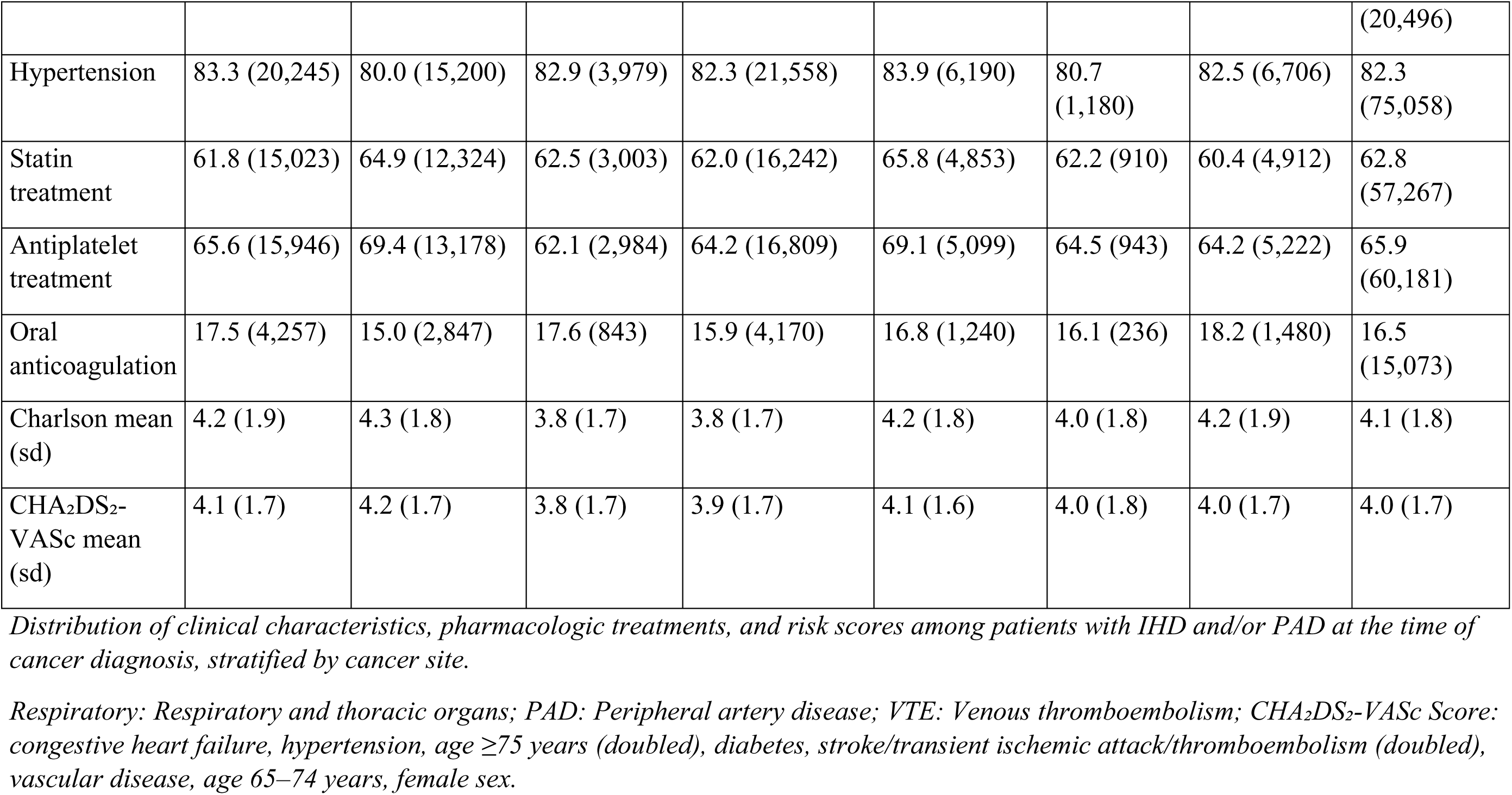
Baseline Demographics, Cardiovascular Comorbidities, and Treatments by Cancer Type.

Comorbidities were frequent, with a mean Charlson Comorbidity Index of 4.1 (SD, 1.8) and a mean CHA₂DS₂-VASc score of 4.0 (SD, 1.7). The most common comorbidities were hypertension (82.3%), diabetes mellitus (22.5%), heart failure (36.5%), atrial fibrillation (20.6%), and prior myocardial infarction (30.1%). Some comorbidities varied by cancer type: stroke was more prevalent among patients with neurologic cancers; PAD was more common in cancers of the respiratory system; and diabetes was more frequent in patients with digestive tract cancers.

Use of cardioprotective medications was high, with 62.8% receiving statins, 65.9% receiving antiplatelet therapy, and 16.5% treated with oral anticoagulants.

Cancer types were distributed as follows: glandular and reproductive organs (28.7%, primarily breast cancer in women and prostate cancer in men), digestive tract (26.6%), respiratory organs (20.8%, mainly lung cancer), hematologic malignancies (8.9%), uro-renal system (8.1%), bone, skin, and connective tissue (5.3%), and neurologic system (1.6%).

### Cardiovascular Outcomes and death

Within 1 year of follow-up, 29,856 patients died, corresponding to an all-cause mortality rate of 42.0 per 100 person-years and a cumulative 1-year mortality of 32.9%. Mortality varied substantially across cancer types (Table 2, Supplementary Figure 1). Patients with cancers of the glandular and reproductive organs had the lowest 1-year mortality (11.8%), whereas those with neurological cancers had the highest (61.8%).

**Table 2.**
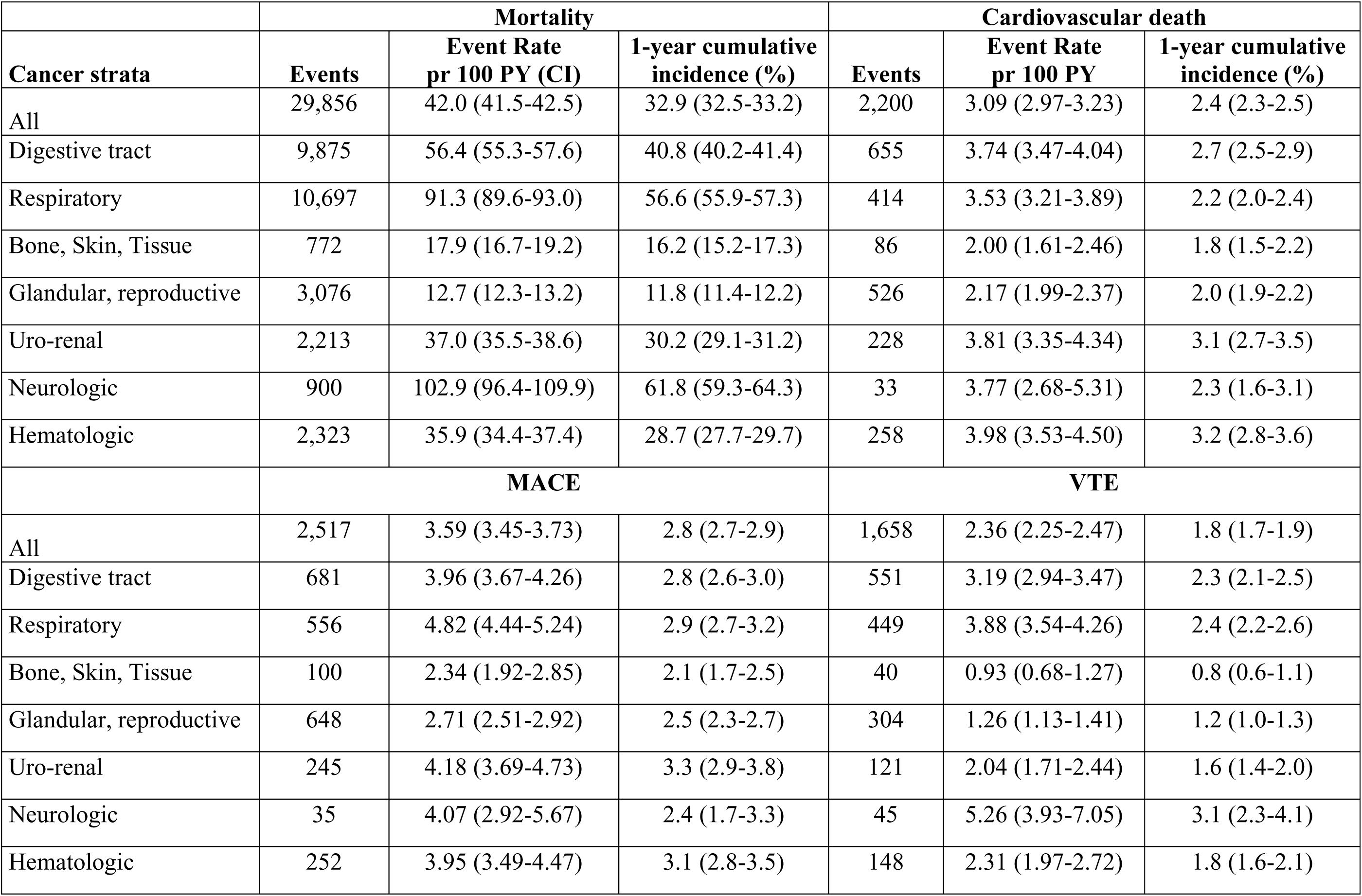
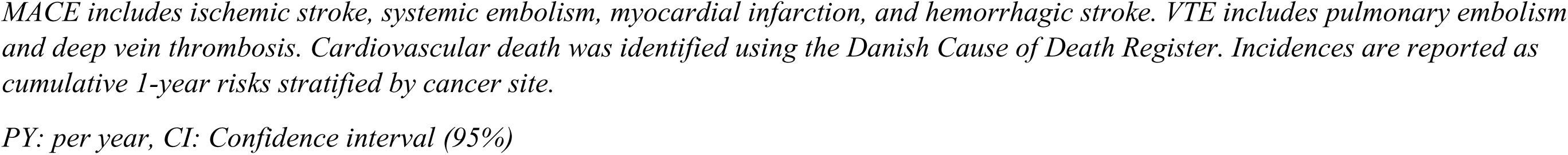
Cardiovascular Outcomes and Mortality by Cancer Type within 1 year of follow-up.

Cardiovascular events occurred at lower rates. A total of 2,200 patients died from cardiovascular causes (3.1 per 100 person-years), 2,517 experienced a major adverse cardiovascular event (MACE; 3.6 per 100 person-years), and 1,658 experienced a VTE event (2.4 per 100 person-years). The corresponding 1-year cumulative incidences were 2.4% for cardiovascular death, 2.8% for MACE, and 1.8% for VTE.

Figure 2 and Table 2 display cumulative incidence curves over the first year following cancer diagnosis, illustrating the timing and burden of cardiovascular events and death. The incidence of MACE and VTE varied by cancer type. MACE was most common among patients with uro-renal cancers (3.3%), and VTE incidence was highest among those with neurologic cancers (3.1%). In contrast, patients with bone, skin, and connective tissue cancers had the lowest incidence of both MACE (2.1%) and VTE (0.8%). Cardiovascular death was most frequent in patients with hematologic (3.2%) and uro-renal (3.1%) malignancies.

**Figure 2.**
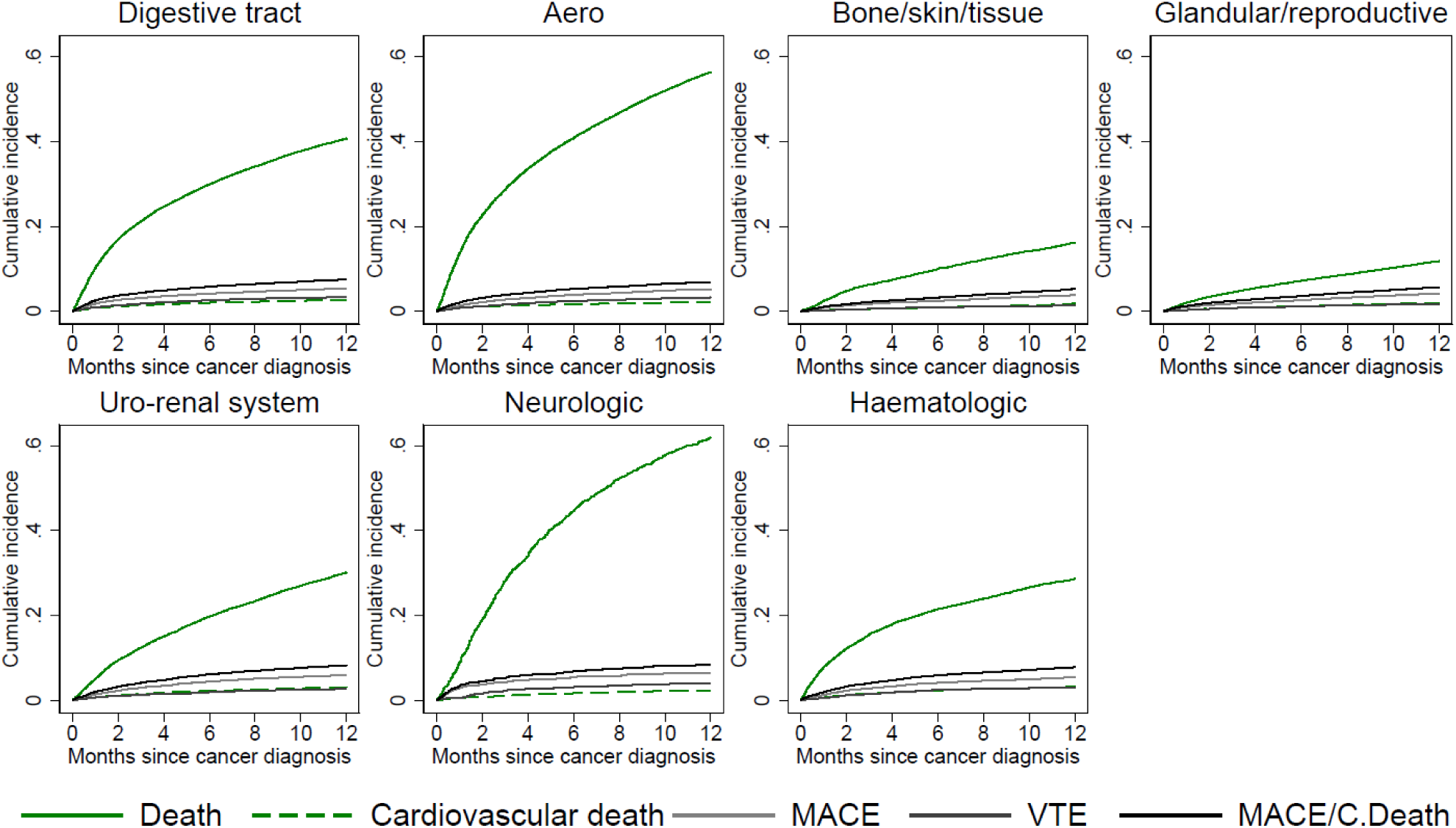
Cumulative 1-Year Incidence of Cardiovascular Events and Death by Cancer Type. *Curves represent cumulative incidence of cardiovascular death, MACE, and VTE over 12 months following cancer diagnosis, stratified by cancer site*. *MACE includes ischemic stroke, systemic embolism, myocardial infarction, and hemorrhagic stroke. VTE includes pulmonary embolism and deep vein thrombosis. Cardiovascular death was identified using the Danish Cause of Death Register. Incidences are reported as cumulative 1-year risks stratified by cancer site*.

### Predictors of cardiovascular outcomes

Crude and adjusted HRs for cardiovascular death, MACE, and VTE were evaluated across a wide range of clinical variables and cancer subtypes. A visual summary of key associations is presented in Figure 3; full estimates are provided in Supplementary Table 2.

**Figure 3.**
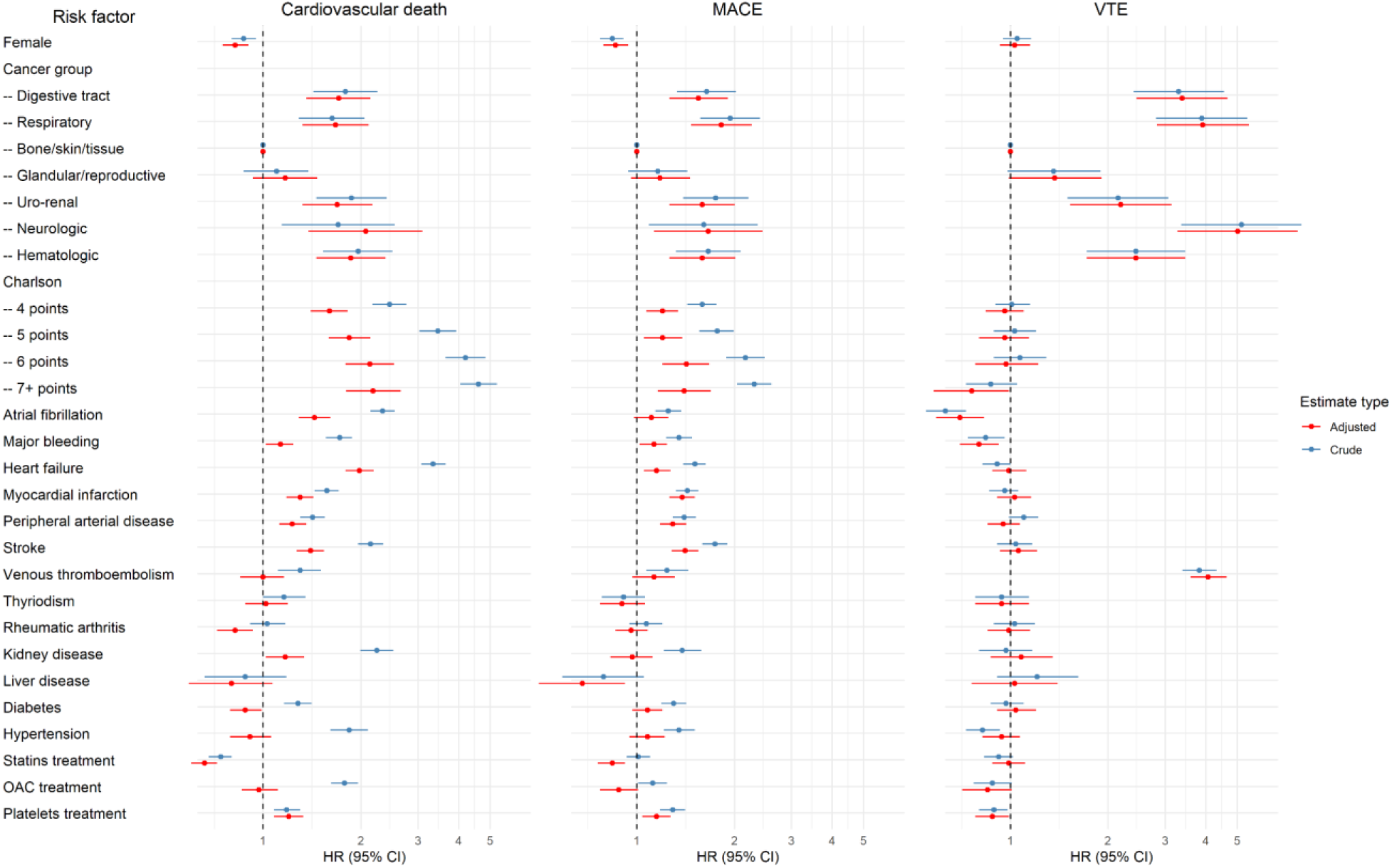
Risk Factors for Cardiovascular Death, MACE and VTE After a Cancer Diagnosis. *Forest plots of hazard ratios (HRs) and 95% confidence intervals for cardiovascular death, MACE, and VTE. Both crude (blue) and adjusted (red) estimates are shown for each risk factor.* *MACE includes ischemic stroke, systemic embolism, myocardial infarction, and hemorrhagic stroke. VTE includes pulmonary embolism and deep vein thrombosis*.

Cardiovascular risk was consistently elevated in the presence of comorbid conditions and varied by cancer type. The strongest associations were observed for neurologic, digestive, and hematologic cancers. Most cardiovascular comorbidities present at the time of cancer diagnosis were associated with increased risk of both cardiovascular death and MACE. Notably, prior stroke (HR, 1.41; 95% CI, 1.28–1.55) and prior myocardial infarction (HR, 1.38; 95% CI, 1.26–1.51) were the most strongly associated with MACE.

Statin use was associated with a significantly reduced risk of cardiovascular death (HR, 0.66; 95% CI, 0.60–0.72) and MACE (HR, 0.84; 95% CI, 0.76–0.92). Similarly, recent use of oral anticoagulants (OAC) showed a tendency toward protective effects for both MACE and VTE. For VTE specifically, atrial fibrillation was associated with a lower risk (HR, 0.70; 95% CI, 0.59–0.83), and prior OAC use was the only medication-related variable linked to increased VTE risk, likely reflecting confounding by indication.

Cancer type remained a strong and independent determinant of cardiovascular outcomes. Compared with cancers of bone, skin, and connective tissue (reference group), all other cancer types—except for glandular and reproductive cancers—were associated with substantially higher risks of cardiovascular death and MACE. A similar pattern was observed for VTE, though effect sizes were generally larger.

These findings highlight the prognostic importance of both baseline comorbidity burden and preventive pharmacotherapy, particularly statins and OAC, in modifying cardiovascular risk after a cancer diagnosis.

## Discussion

The findings from this study highlight that early cardiovascular complications after cancer are not incidental but may reflect interactions between malignancy-related inflammation and pre-existing vascular disease. Patients with IHD or PAD carry a biologically primed vascular environment characterized by endothelial dysfunction, platelet activation, and coagulation pathway upregulation, increasing susceptibility to adverse events.

Although cardiovascular complications are increasingly recognized in cancer care, patients with established vascular disease remain underrepresented in cardio-oncology research and clinical guidelines. In this nationwide cohort of more than 91,000 patients with IHD and/or PAD newly diagnosed with cancer, we found a substantial burden of early mortality and cardiovascular events. Within the first year following cancer diagnosis, nearly one-third of patients died; more than 5 percent experienced a MACE, and approximately 3 percent developed VTE.

The rate of cardiovascular death reached 3 per 100 person-years, underscoring the vulnerability of this dual-disease population. In these patients, systemic atherosclerosis may be further exacerbated by physiological and treatment-related stressors during cancer care.

Marked heterogeneity between cancer groups was observed. This variation is consistent with known differences in tumor-associated inflammation, coagulation pathway activation, metabolic stress, and treatment exposures described in prior literature. These factors may help explain the cardiovascular risk patterns seen across malignancies, although the underlying mechanisms cannot be confirmed within the present dataset. The comparatively higher VTE risk in neurologic cancers and the greater MACE burden in uro-renal cancers may reflect differing vascular risk profiles across malignancies, but these patterns should be interpreted cautiously given the observational design.

### Implications for Vascular Risk Assessment

Cardiovascular risk varied substantially across cancer types. Neurologic, respiratory, and digestive tract cancers were associated with the highest one-year rates of cardiovascular death (Figure 2). Notably, cardiovascular events and cancer-related death often occurred in close temporal proximity, complicating treatment planning and follow-up. Importantly, elevated cardiovascular risk was observed across all cancer groups, extending beyond traditionally recognized “cardiotoxic” malignancies.

In multivariable analyses (Figure 3), several clinical predictors of adverse cardiovascular outcomes emerged consistently. Statin therapy was associated with reduced risk of cardiovascular death (adjusted HR, 0.66), MACE (HR, 0.84), underscoring a potential protective role for lipid-lowering therapy in patients with pre-existing vascular disease undergoing cancer care. The association with VTE risk was neutral (HR, 0.99). This association is biologically plausible given statins’ known effects on plaque stabilization, endothelial function, and systemic inflammation. In contrast, atrial fibrillation, heart failure, stroke, PAD, and higher comorbidity burden were associated with increased cardiovascular risk.

These findings align with prior work emphasizing the prognostic significance of cardiovascular health in oncology.^14,15^ However, most existing studies have focused on therapy-related cardiovascular injury rather than the impact of pre-existing cardiovascular conditions in cancer populations.^16,17^

### Implications for Future Risk Prediction Work

Although large by cardio-oncology standards, our cohort remains too small—and the available data too limited—to support development of a modern, generalizable prediction model. Current guidelines emphasize the need for sufficient event counts, clearly defined clinical use cases, rigorous validation, and transparent reporting.^18^ Even if more detailed data such as biomarkers or imaging parameters were available, many of these inputs would not be accessible at the bedside, limiting practical utility. For a risk tool to be implemented in routine care, it must be automatically populated with information readily available to clinicians.

Pooling all cancer types increases heterogeneity in baseline cardiovascular risk and likely limits model discrimination. Prior studies have shown that models based solely on routinely collected data offer limited predictive performance in oncology populations.^19,20^ While registry-based approaches may support broad population-level stratification, accurate prediction at the individual level will require incorporation of more detailed clinical information. A more feasible next step may be to focus on a single malignancy with shared cardiovascular risk mechanisms—lung cancer is an obvious candidate—and incorporate cancer-specific predictors into risk models.

Recent methodological reviews have cautioned against reliance on simplified sample-size rules and instead recommend formal sample-size calculations, careful modeling of continuous variables, and structured bias assessment, such as PROBAST, to reduce the risk of model failure.^21^

Finally, prognostic models tend to be most reliable when outcomes are relatively infrequent. In high-prevalence settings such as ours, with 30 percent one-year mortality, miscalibration may lead to clinically meaningful misclassification, and predictive values are mathematically constrained. The limited predictive performance of registry-based models underscores that vascular risk in cancer patients is driven by dynamic biological processes, including inflammation, endothelial activation, and hypercoagulability—that are not captured by static clinical variables alone. Before expanding datasets, researchers should clarify the intended clinical use case and determine whether broad risk categorization, rather than precise individualized prediction, may be the more practical strategy for improving patient care.

### Strengths and limitations

To our knowledge, this is the first study to quantify cardiovascular morbidity and mortality in a nationwide real-world cohort of patients with IHD and/or PAD following cancer diagnosis using competing-risk methods and multivariable models. Our findings provide a strong rationale for more targeted research and the development of risk tools tailored to this vulnerable subgroup, which remains underrepresented in trials and underserved in routine care. These results may inform future clinical guidelines and support a shift toward more integrated cardiovascular care in oncology.

Several limitations should be acknowledged. We lacked information on cancer stage, specific oncologic treatments, and relevant biomarkers, all of which may influence cardiovascular risk. The use of administrative codes may have introduced misclassification, although this is likely non-differential. Furthermore, exclusion of metastatic and unspecified cancers may have led to underestimation of cardiovascular risk in advanced disease. Finally, as with all observational studies, residual confounding cannot be excluded.

Future research should bridge population-based vascular epidemiology with mechanistic studies to clarify how cancer perturbs atherosclerotic and thrombotic pathways. Understanding these interactions will be essential for developing targeted vascular interventions during cancer care.

## Conclusions

Early cardiovascular events after cancer diagnosis in patients with IHD or PAD reflect the convergence of malignancy-associated inflammatory and thrombotic stress with advanced vascular disease. Preventive vascular therapies, including statins, may mitigate this risk and should be integrated into oncology care pathways.

## Data Availability

Danish registry data are not publicly available. Access requires formal permission from either Statistics Denmark or the Danish Health Data Authority and can only be granted to approved research institutions. Individual-level data may only be accessed through secure servers and cannot be shared or transferred. This study was conducted under these legal conditions and was approved by the Danish Data Protection Agency (File No. 2012-41-0633).

## Author contributions

All authors had access to the data and a role in writing the manuscript. TBL & FLS: Conceptualization, Visualization, Writing − original draft; FLS: Statistical analysis and data management, MBH, RNHF, CLB: Conceptualization, Validation, Writing − review & editing; TFH: Conceptualization, Writing – review, funding & editing.

## Abbreviations

IHD: ischemic heart disease
PAD: peripheral artery disease
MACE: major adverse cardiovascular events
VTE: venous thromboembolism
ICD-10: International Classification of Diseases, 10th Revision
ATC: Anatomical Therapeutic Chemical
HR: hazard ratio
CI: confidence interval
SD: standard deviation
CHA₂DS₂ VASc: congestive heart failure, hypertension, age ≥75 years (doubled), diabetes, stroke/TIA/thromboembolism (doubled), vascular disease, age 65–74 years, sex category (female)

## References

1. Herrmann J, Lerman A, Sandhu NP, Villarraga HR, Mulvagh SL, Kohli M. Evaluation and management of patients with heart disease and cancer: cardio-oncology. Mayo Clin Proc [Internet]. 2014;89:1287–1306. Available from: http://www.ncbi.nlm.nih.gov/pubmed/25192616

2. Zamorano JL, Lancellotti P, Rodriguez Muñoz D, Aboyans V, Asteggiano R, Galderisi M, Habib G, Lenihan DJ, Lip GYH, Lyon AR, Lopez Fernandez T, Mohty D, Piepoli MF, Tamargo J, Torbicki A, Suter TM, Group ESCSD. 2016 ESC Position Paper on cancer treatments and cardiovascular toxicity developed under the auspices of the ESC Committee for Practice Guidelines: The Task Force for cancer treatments and cardiovascular toxicity of the European Society of Cardiology (ESC. Eur Heart J [Internet]. 2016;37:2768–2801. Available from: http://www.ncbi.nlm.nih.gov/pubmed/27567406

3. Newman AAC, Dalman JM, Moore KJ. Cardiovascular Disease and Cancer: A Dangerous Liaison. Arterioscler Thromb Vasc Biol [Internet]. 2025;45:359–371. Available from: https://www.ahajournals.org/doi/10.1161/ATVBAHA.124.319863

4. Aboumsallem JP, Moslehi J, de Boer RA. Reverse Cardio-Oncology: Cancer Development in Patients With Cardiovascular Disease. J Am Heart Assoc [Internet]. 2020;9. Available from: https://www.ahajournals.org/doi/10.1161/JAHA.119.013754

5. Lyon AR, López-Fernández T, Couch LS, Asteggiano R, Aznar MC, Bergler-Klein J, Boriani G, Cardinale D, Cordoba R, Cosyns B, Cutter DJ, de Azambuja E, de Boer RA, Dent SF, Farmakis D, Gevaert SA, Gorog DA, Herrmann J, Lenihan D, Moslehi J, Moura B, Salinger SS, Stephens R, Suter TM, Szmit S, Tamargo J, Thavendiranathan P, Tocchetti CG, van der Meer P, van der Pal HJH, Group ESCSD. 2022 ESC Guidelines on cardio-oncology developed in collaboration with the European Hematology Association (EHA), the European Society for Therapeutic Radiology and Oncology (ESTRO) and the International Cardio-Oncology Society (IC-OS). Eur Heart J [Internet]. 2022;43:4229–4361. Available from: http://www.ncbi.nlm.nih.gov/pubmed/36017568

6. Wilson PWF, D’Agostino R, Bhatt DL, Eagle K, Pencina MJ, Smith SC, Alberts MJ, Dallongeville J, Goto S, Hirsch AT, Liau C-S, Ohman EM, Röther J, Reid C, Mas J-L, Steg PG, Registry R. An international model to predict recurrent cardiovascular disease. Am J Med [Internet]. 2012;125:695–703.e1. Available from: http://www.ncbi.nlm.nih.gov/pubmed/22727237

7. Dorresteijn JAN, Visseren FLJ, Wassink AMJ, Gondrie MJA, Steyerberg EW, Ridker PM, Cook NR, van der Graaf Y, Group SS. Development and validation of a prediction rule for recurrent vascular events based on a cohort study of patients with arterial disease: the SMART risk score. Heart [Internet]. 2013;99:866–872. Available from: http://www.ncbi.nlm.nih.gov/pubmed/23574971

8. Pedersen CB. The Danish Civil Registration System. Scand J Public Health [Internet]. 2011;39:22–5. Available from: http://www.ncbi.nlm.nih.gov/pubmed/21775345

9. Kildemoes HW, Sørensen HT, Hallas J. The Danish National Prescription Registry. Scand J Public Health [Internet]. 2011 [cited 2012 Dec 9];39:38–41. Available from: http://www.ncbi.nlm.nih.gov/pubmed/21775349

10. Helweg-Larsen K. The Danish Register of Causes of Death. Scand J Public Health [Internet]. 2011;39:26–9. Available from: http://www.ncbi.nlm.nih.gov/pubmed/21775346

11. Albertsen IE, Nielsen PB, Søgaard M, Goldhaber SZ, Overvad TF, Rasmussen LH, Larsen TB. Risk of Recurrent Venous Thromboembolism: A Danish Nationwide Cohort Study. Am J Med [Internet]. 2018;131:1067–1074.e4. Available from: http://www.ncbi.nlm.nih.gov/pubmed/30266273

12. Charlson ME, Pompei P, Ales KL, MacKenzie CR. A new method of classifying prognostic comorbidity in longitudinal studies: development and validation. J Chronic Dis [Internet]. 1987;40:373–83. Available from: http://www.ncbi.nlm.nih.gov/pubmed/3558716

13. Lip GYH, Nieuwlaat R, Pisters R, Lane DA, Crijns HJGM. Refining clinical risk stratification for predicting stroke and thromboembolism in atrial fibrillation using a novel risk factor-based approach: the euro heart survey on atrial fibrillation. Chest [Internet]. 2010 [cited 2012 Oct 7];137:263–72. Available from: http://www.ncbi.nlm.nih.gov/pubmed/19762550

14. Ambrogi V, Pompeo E, Elia S, Pistolese GR, Mineo TC. The impact of cardiovascular comorbidity on the outcome of surgery for stage I and II non-small-cell lung cancer. Eur J Cardiothorac Surg [Internet]. 2003;23:811–817. Available from: http://www.ncbi.nlm.nih.gov/pubmed/12754038

15. Janssen-Heijnen ML, Schipper RM, Razenberg PP, Crommelin MA, Coebergh JW. Prevalence of co-morbidity in lung cancer patients and its relationship with treatment: a population-based study. Lung Cancer [Internet]. 1998;21:105–113. Available from: http://www.ncbi.nlm.nih.gov/pubmed/9829544

16. Florido R, Daya NR, Ndumele CE, Koton S, Russell SD, Prizment A, Blumenthal RS, Matsushita K, Mok Y, Felix AS, Coresh J, Joshu CE, Platz EA, Selvin E. Cardiovascular Disease Risk Among Cancer Survivors: The Atherosclerosis Risk In Communities (ARIC) Study. J Am Coll Cardiol [Internet]. 2022;80:22–32. Available from: http://www.ncbi.nlm.nih.gov/pubmed/35772913

17. Wilcox NS, Amit U, Reibel JB, Berlin E, Howell K, Ky B. Cardiovascular disease and cancer: shared risk factors and mechanisms. Nat Rev Cardiol [Internet]. 2024;21:617–631. Available from: http://www.ncbi.nlm.nih.gov/pubmed/38600368

18. Van Royen FS, Asselbergs FW, Alfonso F, Vardas P, Van Smeden M. Five critical quality criteria for artificial intelligence-based prediction models. Eur Heart J [Internet]. 2023;44:4831–4834. Available from: 10.1093/eurheartj/ehad727

19. Abdel-Qadir H, Amir E, Thavendiranathan P. Prevention, Detection, and Management of Chemotherapy-Related Cardiac Dysfunction. Can J Cardiol [Internet]. 2016;32:891–899. Available from: http://www.ncbi.nlm.nih.gov/pubmed/27118058

20. Kostakou PM, Kouris NT, Kostopoulos VS, Damaskos DS, Olympios CD. Cardio-oncology: a new and developing sector of research and therapy in the field of cardiology. Heart Fail Rev [Internet]. 2019;24:91–100. Available from: http://www.ncbi.nlm.nih.gov/pubmed/30073443

21. Chen L. Overview of clinical prediction models. Ann Transl Med [Internet]. 2020;8:71 Available from: http://atm.amegroups.com/article/view/33251/html

